# Factors Associated with Severe Acute Respiratory Syndrome Coronavirus-2 Infection in Hohoe Municipality, Ghana: A Case-Control Study

**DOI:** 10.1101/2025.09.03.25335047

**Authors:** Sadat Ibrahim, Yussif Yakubu, Kwaku Apiagyei, Adjato Franklin Duncan Sylvester, Yahuza Sabit Tanko, Frank Baiden

## Abstract

Severe acute respiratory syndrome coronavirus-2 (SARS-CoV-2) infection was a major public health challenge globally and in Ghana. To prepare better for future pandemics, an evidence-based understanding of the determinants of the coronavirus disease is essential to inform public health guidelines and surveillance. Thus, we identified the factors for SARS-CoV-2 infection in Hohoe Municipality. We conducted a facility-based, sex and age-matched (1:2) case-control study. Cases were persons with a laboratory-confirmed SARS-CoV-2 infection by Reverse Transcription Polymerase Chain Reaction (RT-PCR) or rapid antigen test, while controls tested negative with the same techniques. Data on sociodemographic, clinical, and exposure-related factors were collected through structured interviews. We employed a conditional regression model to establish the factors independently associated with SARS-CoV-2 infection using Stata version 17.0. All statistical tests were two-sided, and a p-value <0.05 was considered statistically significant. A total of 234 participants were enrolled (78 cases, 156 controls). The mean age of the cases and controls was 39.7±(14.6) and 39.4±(14.4) years, respectively. Moderate/high levels of social interaction increased the odds of infection (aOR=3.00, 95% CI:1.05–8.56, p=0.040). Having no underlying health condition (aOR=0.25, 95% CI:0.09–0.65, p=0.004) and regular physical activity or exercise (aOR=0.18, 95% CI:0.04–0.70, p=0.014) reduced the risk of infection. Moderate/high level of social interaction was associated with increased odds of SARS-CoV-2 infection, and having no underlying condition and frequent exercise/physical activity was protective. Public health interventions should therefore prioritize strengthening community awareness about the risks of close social interactions and the benefits of healthy lifestyles, including regular physical activity.

## Introduction

The novel Severe Acute Respiratory Syndrome Coronavirus-2 (SARS-CoV-2) is the infectious agent of the Coronavirus Disease 2019 (COVID-19) pandemic. It caused widespread infection, morbidity, and mortality since late 2019 and created a global health emergency *[1]*. On January 30, 2020, the World Health Organisation (WHO) declared SARS-CoV-2 a public health emergency of international concern, and a pandemic on March 11, 2020 *[1,2]*. Ghana’s index case was confirmed on March 12, 2020 *[3]*.

Due to insufficient testing capacity and a high proportion of asymptomatic infection, the pandemic swiftly changed, resulting in widespread under-detection and many unaccounted cases *[4,5]*. Despite the possibility of symptomatic disease, a significant proportion of the population were asymptomatic carriers, and clinical presentations ranged from mild to severe or critical *[6]*. Over 80% of the cases were asymptomatic or presented with mild symptoms, implying a lot of asymptomatic incubatory and healthy carriers *[2].* This made symptomatic patients a hidden potential source of transmission. Direct or indirect contact with respiratory aerosols, droplets or other bodily fluids via contaminated surfaces is the known predominant mode of transmission *[7,8]*.

The Centers for Disease Control and Prevention defined risk factors as a variety of factors that could influence an individual’s exposure, or response to a causative agent and increase susceptibility, or predispose to a disease or infection *[9]*. Although COVID-19 is mainly transmitted through direct or indirect exposure to the etiologic agent, transmission dynamics have identified demographics like advanced age, clinical profiles or cardiovascular diseases, including hypertension and diabetes, and travel history to predispose to the infection and may account for severe outcomes *[10,11]*. Moreover, environmental factors like temperature and humidity, health system organisation and policy, politico-economic situation, large gatherings, and international travel could substantially drive community transmission *[12,13,14]*

Even with the abundance of epidemiological data produced during the pandemic, significant knowledge gaps persist regarding the specific factors associated with SARS-CoV-2 infection in rural and peri-urban African communities. Thus far, no prior studies have established evidence on the determinants of SARS-CoV-2 infection within Hohoe Municipality. We identified the factors for SARS-CoV-2 infection in Hohoe Municipality. We seek to provide evidence-based insights that can guide surveillance and local public health interventions and contribute to the broader understanding of SARS-CoV-2 infection epidemiology in the study setting and Ghana by extension. The results contribute to the larger body of global health evidence on SARS-CoV-2 disease. Also, it offers implications for informing public health efforts, strengthening surveillance and pandemic preparedness and response to emerging and reemerging infectious illnesses.

## Methods

### Study Design

We adopted a matched case-control design to identify the factors for SARS-CoV-2 infection. To eliminate confounders, the cases were individually matched (1 case: 2 controls) to the controls, in which the enrolled controls were identical to the cases in age (*±5 years*) and sex.

### Study Site

Hohoe Municipality, with Hohoe as the capital, is where the study was conducted. It was carved out of the Kpando District and borders the Republic of Togo on the east, on the southeast by the Afadzato district and southwest by the Kpando Municipality; on the north with the Guan District, and on the northwest with the Biakoye District. The municipality has a population of 114,472, representing 6.8% of the region’s total population *[15]*. It comprises 52.1% females and 47.9% males. About fifty-three percent of the population is urban and comprises eleven major towns/settlements *[15]*. Regarding healthcare, the municipality has four sub-districts, the Volta Regional Hospital, six community Health-Based Planning Services Compounds, and eight health centers. The Volta Regional Hospital is the municipality’s largest public health and referral facility.

### Study Population

The study was among persons resident in Hohoe Municipality who tested for SARS-CoV-2 by Reverse transcription polymerase chain reaction (RT-PCR) or rapid antigen test between March 2020 and December 2021. If the test result was positive, the person was classified as a case; otherwise, as a potential control. Persons aged 18 years or above, and with a positive or negative laboratory-confirmed diagnosis of SARS-CoV-2 by RT-PCR or antigen test between March 2020 and December 2021 were considered for inclusion. The exclusion criteria were persons who tested positive for SARS-CoV-2 other than an RT-PCR or rapid antigen test, tested positive later than December 2021, had previously tested positive for SARS-CoV-2 and subsequently tested positive again (as potential cases of reinfection), had travelled at the time of data collection, had debilitating health conditions or poor cognitive ability or incomplete or missing relevant information in the SARS-CoV-2 infection-line list.

### Sample Size

A priori statistical power calculation using Fleiss with Continuity Correction approach *[16]* was used to determine the study’s sample size, 237 (79 cases, 158 controls).

### Sampling Method

We employed systematic sampling to recruit the cases using the line list of the cases as a sample frame. The 79 cases were calculated proportionate to the 506 eligible cases on the line list. The sampling interval (6^th^ term) was determined by dividing the total number of cases (506) by the sample size of the cases (79). Having determined this, the first case was randomly selected between the first case and the 6^th^ term on the line list. For the controls, the cases were individually matched (1 case: 2 controls) to the controls by age (±5 years) and sex.

### Data Collection Procedure

A standard structured questionnaire was used to gather data between September 2, 2023 and October 28, 2023. A face-to-face interview was conducted, and data were captured electronically with KoboCollect. Written informed consent was obtained from the individual respondents before the questionnaire was administered.

### Study Variables

Age, sex, educational level, occupation, marital status, income level, ethnicity, and residence were used to gather sociodemographic data. Clinical information, SARS-CoV-2 infection contact history, infection exposure and outcomes were also collected, including **s**elf-reported risk information on alcohol use, and smoking history/status.

### Statistical Analyses

Data collated on KoboCollect was extracted in Microsoft Excel format. We then checked and resolved all discrepant data before formal analyses using Stata version 17.0 statistical software. Categorical variables were described using frequencies and percentages, and the mean (standard deviation) was computed for continuous variables. Given the individually age- and sex-matched (1:2) case–control design, associations between potential risk factors and SARS-CoV-2 infection were first explored using univariate conditional logistic regression, reporting crude odds ratios (cORs) with 95% confidence intervals (CIs). Variables with a p-value <0.20 in the univariate analyses were considered for inclusion in a multivariable conditional logistic regression model. Adjusted odds ratios (aORs) with 95% CIs were then estimated to identify factors independently associated with SARS-CoV-2 infection. All statistical tests were two-sided, and a p-value <0.05 was considered statistically significant

## Results

### Sociodemographic characteristics of cases and controls

Data was gathered from 234 respondents and the mean ages of the cases and controls were 39.7± (14.6) and 39.4± (14.4) years, respectively with an overall mean age of 39.5± (14.5) years (Table 1). Urban residence was higher among cases 62 (79.5%) versus controls 94 (60.3%). Less than half of the cases 21 (26.9%) were asymptomatic as against the controls 89 (57.0%). Of the cases, less than half 33 (42.3%) had been prediagnosed with a health condition compared to the controls 108 (69.2%). The proportion of cases with hypertension and diabetes was 23 (29.5%) and 18 (23.1%), respectively; versus hypertension 23 (14.7%) and diabetes 21 (13.5%) in the control group.

**Table 1:**
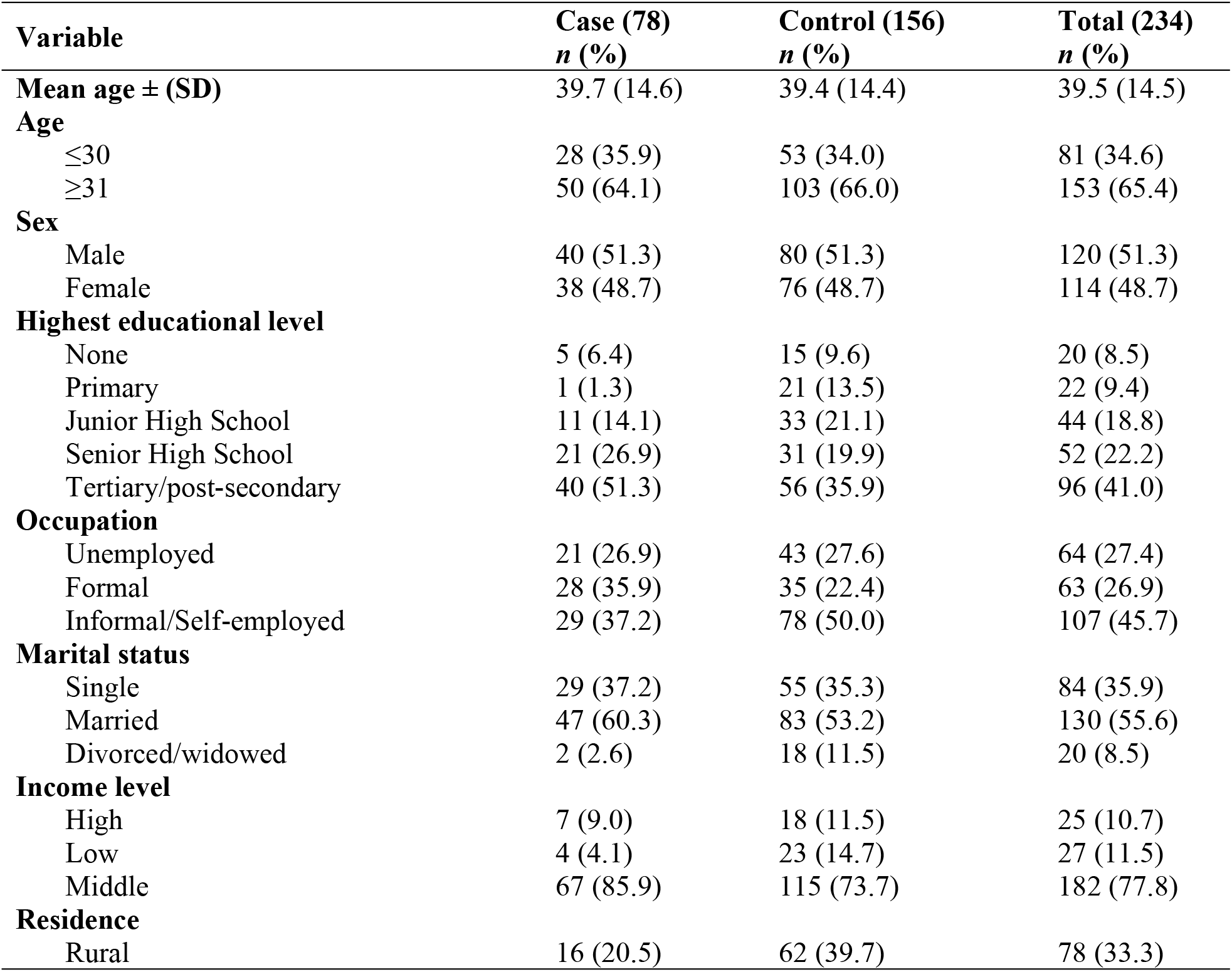
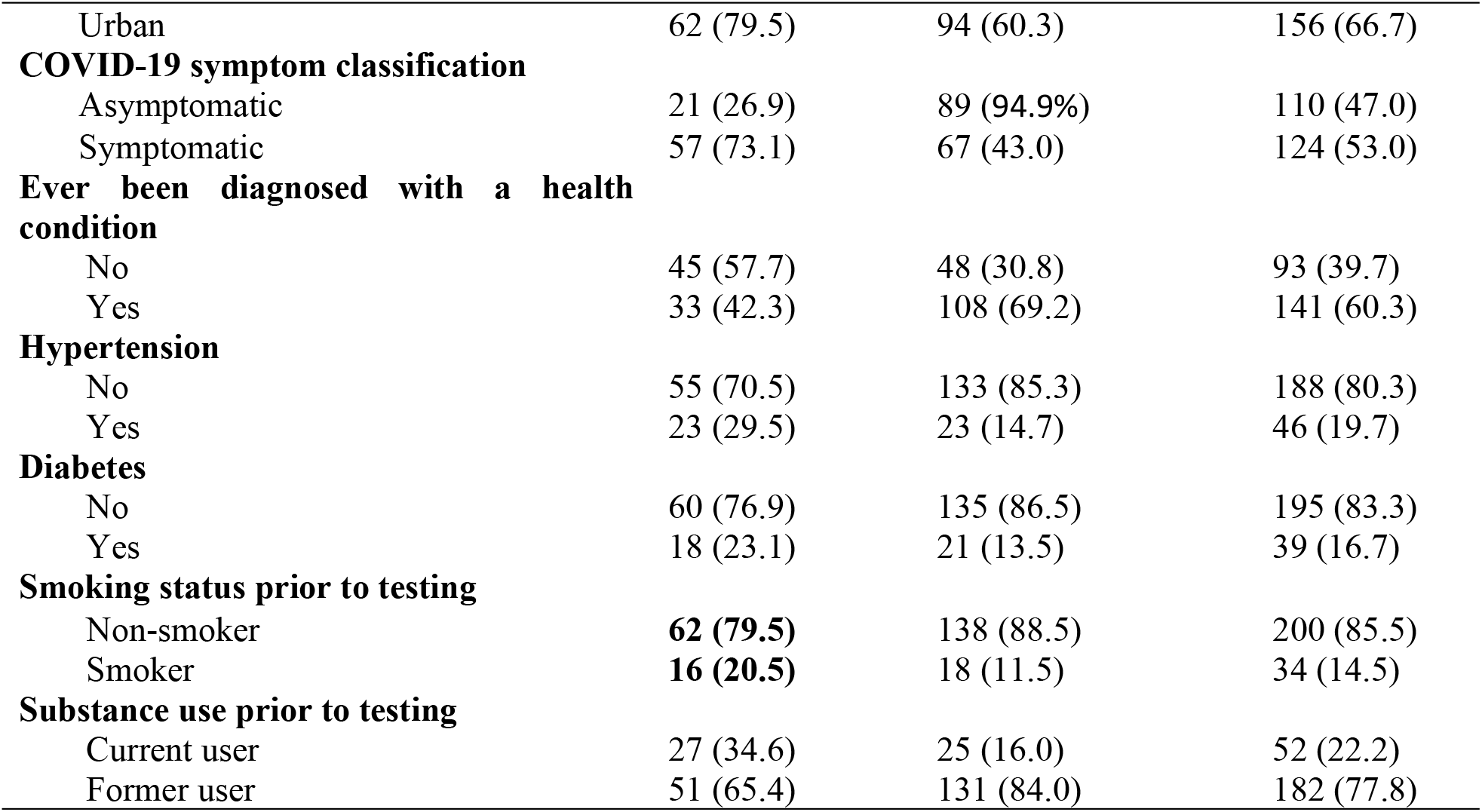
Sociodemographic characteristics of cases and controls.

| Variable | Case (78)<br>n (%) | Control (156)<br>n (%) | Total (234)<br>n (%) |
| --- | --- | --- | --- |
| **Mean age ± (SD)** | 39.7 (14.6) | 39.4 (14.4) | 39.5 (14.5) |
| **Age** |  |  |  |
| ≤30 | 28 (35.9) | 53 (34.0) | 81 (34.6) |
| ≥31 | 50 (64.1) | 103 (66.0) | 153 (65.4) |
| **Sex** |  |  |  |
| Male | 40 (51.3) | 80 (51.3) | 120 (51.3) |
| Female | 38 (48.7) | 76 (48.7) | 114 (48.7) |
| **Highest educational level** |  |  |  |
| None | 5 (6.4) | 15 (9.6) | 20 (8.5) |
| Primary | 1 (1.3) | 21 (13.5) | 22 (9.4) |
| Junior High School | 11 (14.1) | 33 (21.1) | 44 (18.8) |
| Senior High School | 21 (26.9) | 31 (19.9) | 52 (22.2) |
| Tertiary/post-secondary | 40 (51.3) | 56 (35.9) | 96 (41.0) |
| **Occupation** |  |  |  |
| Unemployed | 21 (26.9) | 43 (27.6) | 64 (27.4) |
| Formal | 28 (35.9) | 35 (22.4) | 63 (26.9) |
| Informal/Self-employed | 29 (37.2) | 78 (50.0) | 107 (45.7) |
| **Marital status** |  |  |  |
| Single | 29 (37.2) | 55 (35.3) | 84 (35.9) |
| Married | 47 (60.3) | 83 (53.2) | 130 (55.6) |
| Divorced/widowed | 2 (2.6) | 18 (11.5) | 20 (8.5) |
| **Income level** |  |  |  |
| High | 7 (9.0) | 18 (11.5) | 25 (10.7) |
| Low | 4 (4.1) | 23 (14.7) | 27 (11.5) |
| Middle | 67 (85.9) | 115 (73.7) | 182 (77.8) |
| **Residence** |  |  |  |
| Rural | 16 (20.5) | 62 (39.7) | 78 (33.3) |
| Urban | 62 (79.5) | 94 (60.3) | 156 (66.7) |
| **COVID-19 symptom classification** |  |  |  |
| Asymptomatic | 21 (26.9) | 89 (94.9%) | 110 (47.0) |
| Symptomatic | 57 (73.1) | 67 (43.0) | 124 (53.0) |
| **Ever been diagnosed with a health condition** |  |  |  |
| No | 45 (57.7) | 48 (30.8) | 93 (39.7) |
| Yes | 33 (42.3) | 108 (69.2) | 141 (60.3) |
| **Hypertension** |  |  |  |
| No | 55 (70.5) | 133 (85.3) | 188 (80.3) |
| Yes | 23 (29.5) | 23 (14.7) | 46 (19.7) |
| **Diabetes** |  |  |  |
| No | 60 (76.9) | 135 (86.5) | 195 (83.3) |
| Yes | 18 (23.1) | 21 (13.5) | 39 (16.7) |
| **Smoking status prior to testing** |  |  |  |
| Non-smoker | **62 (79.5)** | 138 (88.5) | 200 (85.5) |
| Smoker | **16 (20.5)** | 18 (11.5) | 34 (14.5) |
| **Substance use prior to testing** |  |  |  |
| Current user | 27 (34.6) | 25 (16.0) | 52 (22.2) |
| Former user | 51 (65.4) | 131 (84.0) | 182 (77.8) |

### Contact History and Disease Exposure of Cases and Controls

Table 2 illustrates the contact history and disease exposure of the respondents. Among the cases, the majority 47 (60.3%) attended a gathering of less than ten people, and a greater 54 (69.2%) proportion travelled within Ghana. Most 65 (83.3%) of the cases had a moderate/high level of frequent social interactions compared to 82 (52.6%) of the controls.

**Table 2:** Contact History and Disease Exposure of Cases and Controls.

| Variable | Case (78)<br>*n* (%) | Control (156)<br>*n* (%) | Total (234)<br>*n* (%) |
| --- | --- | --- | --- |
| **Lived in a densely populated or crowded area prior to testing** |  |  |  |
| No | 55 (70.5) | 123 (78.8) | 178 (76.1) |
| Yes | 23 (29.5) | 33 (21.2) | 56 (23.9) |
| **Attended a mass gathering prior to testing** |  |  |  |
| No | 43 (55.1) | 95 (60.9) | 138 (59.0) |
| Yes | 35 (44.9) | 61 (39.1) | 96 (41.0) |
| **Attended a gathering of fewer than ten people prior to testing** |  |  |  |
| No | 31 (39.7) | 92 (59.0) | 123 (52.6) |
| Yes | 47 (60.3) | 64 (41.0) | 111 (47.4) |
| **Travelled to a place within Ghana** |  |  |  |
| No | 24 (30.8) | 98 (62.8) | 122 (52.1) |
| Yes | 54 (69.2) | 58 (37.2) | 112 (47.9) |
| **Travel location** |  |  |  |
| Rural | 14 (25.9) | 25 (43.9) | 39 (35.1) |
| Urban | 40 (74.1) | 32 (56.1) | 72 (64.9) |
| **Visited a healthcare facility prior to testing** |  |  |  |
| No | 34 (43.6) | 103 (66.0) | 137 (58.6) |
| Yes | 44 (56.4) | 53 (34.0) | 97 (41.4) |
| **Had close contact with a suspected or confirmed COVID-19 case prior to testing** |  |  |  |
| No | 63 (80.8) | 146 (93.6) | 209 (89.3) |
| Yes | 15 (19.2) | 10 (6.4) | 25 (10.7) |
| **Had close contact with a generally ill person prior to testing** |  |  |  |
| No | 58 (74.4) | 146 (93.6) | 204 (87.2) |
| Yes | 20 (25.6) | 10 (6.4) | 30 (12.8) |
| **Usual/most frequent means of transportation prior to testing** |  |  |  |
| Private means | 18 (23.1) | 53 (34.0) | 71 (30.3) |
| Public means | 60 (76.9) | 103 (66.0) | 163 (69.7) |
| **Level of frequent social interactions prior to testing** |  |  |  |
| Low crowding | 13 (16.7) | 82 (52.6) | 95 (40.6) |
| Moderate/high crowding | 65 (83.3) | 74 (47.4) | 139 (59.4) |
| **Place of most possible exposure** |  |  |  |
| **Health facility** |  |  |  |
| No | 59 (75.6) | 146 (93.9) | 205 (87.6) |
| Yes | 19 (24.4) | 10 (6.4) | 29 (12.4) |
| **Workplace** |  |  |  |
| No | 59 (75.6) | 111 (71.2) | 170 (72.6) |
| Yes | 19 (24.4) | 45 (28.8) | 64 (27.4) |
| **Had access to PPE** |  |  |  |
| No | 30 (38.5) | 56 (35.9) | 86 (36.7) |
| Yes | 48 (61.5) | 100 (64.1) | 148 (63.3) |

**Table 3:** Univariate and multivariate logistic regression analyses of predictors for SARS-CoV-2 Infection.

| Variable |  | cOR (95% CI) | *p-value* | aOR (95% CI) | *p-value* |
| --- | --- | --- | --- | --- | --- |
| **Residence** |  |  |  |  |  |
|  | Rural | Ref. |  | Ref. |  |
|  | Urban | **2.44 (1.30-4.60)** | **0.006** | 1.16 (0.45-3.03) | 0.758 |
| **Household residence type** |  |  |  |  |  |
|  | Compound house setting | Ref. |  | Ref. |  |
|  | Private/self-contained | **0.55 (0.31-0.95)** | **0.032** | 0.57 (0.25-1.30) | 0.182 |
| **National Health Insurance Scheme (NHIS) status prior to testing** |  |  |  |  |  |
|  | Inactive | Ref. |  | Ref. |  |
|  | Active | **1.93 (1.00-3.74)** | **0.050** | 1.51 (0.52-4.37) | 0.447 |
| **Symptom classification** |  |  |  |  |  |
|  | Asymptomatic | Ref. |  | Ref. |  |
|  | Symptomatic | **3.57 (1.92-6.63)** | **<0.001** | 2.21 (0.90-5.41) | 0.082 |
| **Number of children had prior to testing** |  |  |  |  |  |
|  | None | Ref. |  | Ref. |  |
|  | ≥1 | **0.33 (0.11-0.96)** | **0.042** | 0.21 (0.03-1.37) | 0.103 |
| **Diagnosed with underlying health condition(s) prior to testing** |  |  |  |  |  |
|  | No | **0.25 (0.13-0.50)** | **<0.001** | **0.25 (0.09-0.65)** | **0.004** |
|  | Yes | Ref. |  | Ref. |  |
| **Engaged in physical activities or exercises regularly prior to testing** |  |  |  |  |  |
|  | No | Ref. |  | Ref. |  |
|  | Yes | **0.17 (0.06-0.45)** | **<0.004** | **0.18 (0.04-0.70)** | **0.014** |
| **Attended gathering with less than ten people prior to testing** |  |  |  |  |  |
|  | No | Ref. |  | Ref. |  |
|  | Yes | **2.13 (1.22-3.71)** | **0.008** | 2.13 (0.87-5.18) | 0.097 |
| **Travelled to another region, town or place prior to testing** |  |  |  |  |  |
|  | No | Ref. |  | Ref. |  |
|  | Yes | **3.42 (1.92-6.07)** | **<0.007** | 2.17 (0.92-5.10) | 0.076 |
| **Visited a healthcare facility prior to testing** |  |  |  |  |  |
|  | No | Ref. |  | Ref. |  |
|  | Yes | **2.57 (1.44-4.58)** | **0.001** | 1.86 (0.73-4.76) | 0.195 |
| **Had close contact with a suspected or confirmed COVID-19 case prior to testing** |  |  |  |  |  |
|  | No | Ref. |  | Ref. |  |
|  | Yes | **3.72 (1.50-9.24)** | **0.005** | 0.64 (0.12-3.33) | 0.598 |
| **Had close contact with a generally ill person prior to testing** |  |  |  |  |  |
|  | No | Ref. |  | Ref. |  |
|  | Yes | **4.30 (1.95-9.46)** | **<0.001** | 1.16 (0.30-4.43) | 0.832 |
| **Level of frequent social interactions prior to testing** |  |  |  |  |  |
|  | Low crowding | Ref. |  | Ref. |  |
|  | Moderate/high crowding | **5.82 (2.80-12.11)** | **<0.001** | **3.00 (1.05-8.56)** | **0.040** |
| **Health facility** |  |  |  |  |  |
|  | No | Ref. |  | Ref. |  |
|  | Yes | **4.85 (2.02-11.63)** | **<0.001** | 1.51 (0.26-8.65) | 0.645 |

*aOR – Adjusted Odds Ratio, cOR – Crude Odds Ratio, CI – Confidence Interval*
*\*Boldface type indicates a statistically significant difference at $p < 0.05$*

### Univariate and Conditional Multivariate Logistic Regression Analyses of Severe Acute Respiratory Syndrome Coronavirus-2 Infection Risk Factors

Univariate logistic regression analysis displayed in Table 4 revealed several factors statistically associated with SARS-CoV-2 infection. These included household residence type (cOR=0.55, 95% CI: 0.31-0.95, p=0.032), symptom classification (cOR=3.57, 95% CI: 1.92-6.63, p<0.001), number of children (cOR=0.33, 95% CI: 0.11-0.96, p=0.042), physical activity (cOR=0.17, 95% CI: 0.06-0.45, p<0.004), gathering of less than ten people (cOR=2.13, 95% CI: 1.22-3.71, p<0.008), and level of social interaction (cOR=5.82, 95% CI: 2.80-12.11, p<0.001). Additionally, a health facility as a possible place of exposure (cOR=4.85, 95% CI: 2.02-11.63, p<0.001) showed an association. NHIS status (cOR=1.93, 95% CI: 1.00-3.74, p=0.050) was borderline statistically significant.

In the conditional multivariate model, moderate/high social interaction was associated with higher odds of SARS-CoV-2 infection (aOR 3.00, 95% CI 1.05–8.56). Absence of an underlying condition (aOR 0.25, 95% CI 0.09–0.65) and regular physical activity (aOR 0.18, 95% CI 0.04–0.70) were associated with lower odds. Associations for attending small gatherings and recent travel were elevated in crude analyses but were not statistically significant after adjustment.

## Discussion

Numerous studies have demonstrated factors that facilitate susceptibility or predisposition to COVID-19. There is a clear link between certain pre-existing health conditions and SARS-CoV-2 infection and its adverse prognosis *[17].* This study found hypertension and diabetes to be the prevalent comorbid conditions, which are not different from findings reported by *[18,19,20,21].* Our study showed a syndemic relationship between SARS-CoV-2 infection and preexisting health conditions. A similar study *[21]* demonstrated that comorbidity emerged statistically significant with SARS-CoV-2 infection. Additionally, in support, *[22]* reported that SARS-CoV-2 infection was predominant among people who presented with a comorbidity. Furthermore, the outcome of a national study in Saudi Arabia disclosed that a high proportion of the COVID-19 patients had a history of at least one lifelong illness, which seemingly supports this study’s finding *[23].* On account of the evidence from the above studies, whose findings corroborate that of our study, it is imperative to consider and tailor actions that will safeguard the health needs of persons with a preexisting diagnosis. Individuals with a longstanding illness tend to face severe or critical clinical outcomes or even death. Our findings imply that further studies are vital to assess the clinical outcomes of SARS-CoV-2 infection cases in the study jurisdiction.

Epidemiological link is crucial to understanding SARS-CoV-2 infection transmission dynamics, spread and evidence-based control actions. The current study established that attendance at small gatherings showed an increased risk, but was however, not statistically significant in the in the final multivariate model. Inversely, this finding is on par with *[18,24]* which reported that attending a mass social gathering was associated with SARS-CoV-2 infection. Small gatherings create a perceived sense of safety leading to more relaxed precautions and preventive behaviors such as mask use, physical distancing and attention to ventilation. This false sense of security could facilitate transmission. Moreover, such gatherings typically occur in indoor settings where airflow is limited and individuals remain in proximity for extended periods. In contrast, larger gatherings, which have been more strictly regulated or subject to formal preventive protocols during the pandemic may have entailed greater adherence to public health guidelines.

Regular physical activity has been associated with enhanced immune function, which contributes to a strengthened immune system, potentially reducing the severity and duration of SARS-CoV-2 infection *[25,26].* Our study uncovered that regular exercise or physical activity decreased the risk of SARS-CoV-2 infection. A similar finding reported by *[21]* indicated that physical exercise or activity lessened the odds of infection. In support of the above assertions, a study in the United States revealed a negative correlation between physical activity and SARS-CoV-2 infection further showing a worse prognosis of death among those whose physical activity was less. *[27].* Moreover, according to *[28],* adults between the ages of 40 and 69, those who were physically active were less likely to be confirmed SARS-CoV-2 seropositive. However, in dissonance with the findings aforementioned, *[29]* found that physical activity was not associated with the risk of SARS-CoV-2 infection. Similarly, it was also conversely reported that physical activity had no protective association between SARS-CoV-2 infection and its related symptoms [30]. Physical inactivity predisposes to longstanding diseases like hypertension and diabetes, which are established comorbidities that increase SARS-CoV-2-infection susceptibility or severity and point out the significance of regular physical activity and exercise in SARS-CoV-2 infection prevention *[30].* Although it is widely asserted that regular physical activity is a non-pharmacological intervention against SARS-CoV-2 infection and could help alleviate the symptoms and severity *[31],* there is uncertainty regarding the effects on SARS-CoV-2-infected individuals, despite some evidence to the effect that physical exercise improves respiratory and physical health.

### Strengths and Limitations

This study provides localised evidence for guiding tailored public health interventions and contributes to the broader understanding of SARS-CoV-2 epidemiology. A major strength of this study is the use of a matched case-control design, which effectively controlled for major confounders, thereby enhancing the internal validity of the identified associations. Nonetheless, certain limitations must be acknowledged. A notable limitation was the potential of recall bias which may have stemmed from respondents’ inability to accurately recall or report their exposure history or conditions, as the study was carried out long after the pandemic (time lag between exposure (2020–2021) and data collection (2023). Second, the study might have been prone to social desirability bias in the reporting of preventive behaviors. Particularly, cases may have overestimated adherence to preventive behaviors and protocols which could result in weak or reverse true associations. However, the use of a standard and validated closed-ended questionnaire was used to improve respondents’ information. Lastly, missing information such as contact address in the line list records may have introduced some selection bias during sampling.

## Conclusion

This study identified moderate to high levels of social interaction as the main risk factor for SARS-CoV-2 infection, while regular physical activity and the absence of preexisting health conditions were protective. These findings reinforce the importance of maintaining safe social behaviors during outbreaks and highlight the role of individual health status in susceptibility to infection. Public health interventions should therefore prioritize strengthening community awareness about the risks of close social interactions and the benefits of healthy lifestyles, including regular physical activity. Moreover, tailored strategies should be developed to protect individuals with underlying health conditions, who remain particularly vulnerable. Lessons from this study provide important insights for strengthening preparedness and response strategies against future pandemics in Ghana and similar settings. Further research is needed to explore the long-term effects of these risk and protective factors on SARS-CoV-2 infection outcomes.

## Data Availability

We have the data and would like the journal to help us make it available in any of the repositories.

## Acknowledgement

The authors gratefully acknowledge the Hohoe Municipal Director of Health Services, the Disease Control Officers of the Hohoe District Health Directorate and all individuals whose cooperation and invaluable contributions in various capacities made this work possible.

## Authors’ contributions

Conceptualization and writing official draft: S.I., and F.B., Data collection: S.I., Y.Y., Y.S.T., and K.A, Data Curation and formal Analyses: S.I., A.F.D.S, Y.S.T and Y.Y., Editing and final review: F.B., Y.Y., K.A. All authors read and approved the current version of the manuscript.

### Abbreviations

COVID-19: Coronavirus Disease
RT-PCR: Reverse transcription polymerase chain reaction
SARS-CoV-2: Severe Acute Respiratory Syndrome Coronavirus-2
WHO: World Health Organization

## Declarations

### Availability of data and materials

The anonymized dataset will be deposited in an open repository.

### Ethical Considerations

The study protocol was approved by the University of Health and Allied Sciences Research Ethics Committee (approval ID: *UHAS-RECA.8[88]22-23*) and the Ghana Health Service Ethics Review Committee (approval ID: *GHS-ERC:029/06/23*).

